# Comparison of 2D and 3D oxygen-enhanced MRI of the placenta

**DOI:** 10.1101/2023.03.17.23287353

**Authors:** Penny L. Hubbard Cristinacce, Minal Patel, Alexander Oh, Josephine H. Naish, Edward D. Johnstone, Emma Ingram

## Abstract

Oxygen-Enhanced Magnetic Resonance Imaging (OE-MRI) of the human placenta is potentially a sensitive marker of in vivo oxygenation. This methodological study shows that full coverage of the placenta is possible using 3D mapping of the change in longitudinal relaxation rate (ΔR_1_), in a group of healthy pregnant subjects breathing elevated levels of oxygen.

Twelve pregnant subjects underwent a comparison of 2D and 3D OE-MRI. ΔR_1_ was mapped for a single 2D slice (ss-2D), a single matched-slice from the 3D volume (ss-3D) and the full 3D volume (vol-3D).

The group-average median ΔR_1_ values for ss-3D (0.023 s^-1^) and vol-3D (0.022 s^-1^) do not differ significantly from ss-2D (0.020 s^-1^), when compared using a two-tailed paired t-test (ss-3D (p = 0.58) and vol-3D (p = 0.70)). However, median baseline T_1_ (T_1b_) for ss-2D was higher (1603 ms) than T_1b_ for ss-3D (1540 ms, p=0.07) and significantly higher than vol-3D (1515 ms, p = 0.02), when compared using a two-tailed paired t-test. In contrast with previous studies, no correlation of median ΔR_1_ with gestation age at scan for the normal group (N = 10) was observed for ss-2D, likely due to the smaller gestational range.

Full volume OE-MRI maps reveal sensitivity to changes in ΔR_1_, with some participants showing an enhanced gradient in the intermediate space between the fetal and maternal sides of the placenta in the 3D data. This study shows that it is feasible to acquire whole placental volume OE-MRI data on a Philips 1.5 T MRI scanner. Further work is necessary to implement this increased coverage on the scanners of other vendors.

## Introduction

Oxygen-Enhanced Magnetic Resonance Imaging (OE-MRI) of the human placenta has shown promise in examining placental function(1,2) and dysfunction(3) by being sensitive to in vivo oxygenation. Abnormalities of placental function can be associated with fetal growth restriction (FGR), which leads to increased neonatal morbidity and mortality(4,5). Non-invasive measurements of placental hypoxia, which is associated with peripheral hypovascularity and increased vascular resistance(6), have the potential to identify FGR independent of fetal size.

The spin-lattice relaxation time (T_1_) of tissue is shortened when a subject breathes an elevated concentration of oxygen due to the paramagnetic effect of the additional dissolved oxygen. This change can be quantitatively assessed by examining the change in R_1_ (where R_1_ = 1/T_1_), when a subject is switched from breathing air to 100% oxygen. Previous studies(1–3) have investigated this change by acquiring a single two-dimensional (2D) T_1_-weighted slice through the placenta. Here we investigate the feasibility of obtaining whole placental oxygenation maps using a three-dimensional (3D) volume acquisition.

The aim of the study was to investigate whether a 3D volume acquisition shows the same quantitative trends in R_1_ as a single 2D slice, and to assess whether the increased volume covered could offer more sensitivity to placental heterogeneity. Full 3D coverage of the placenta may therefore allow a more comprehensive assessment of the placental oxygenation, as well as assessing known changes in gross tissue morphology.

## Methods

Twelve subjects were recruited from St Mary’s Hospital, Manchester, following informed consent (ethical approval REC:09/H1013/77 and 14/NW/0195). A summary of subject demographic and pregnancy outcome is given in Table 1. All subjects had normal uterine artery Doppler flow (pulsatility index <95^th^ centile at 22-24 weeks) and normal umbilical artery Doppler flow (pulsatility index <95th percentile and positive end-diastolic flow)(9). Pregnancies were subsequently classified as potentially abnormal based on an individualised birth ratio (IBR) <5^th^ percentile (Gestation-related Optimal Weight (10)). Two women did not meet the outcome criteria for assignment to the normal group.

**Table 1.** Subject demographic and pregnancy outcomes. *Abnormal based on an individualised birth ratio (IBR) <5^th^ percentile.

| Participant No. | Maternal Age Range (years) | BMI | Gestational age at scan (days) | Placental position | Delivery Gestation (days) | Birth weight (g) | IBR (centile) |
| --- | --- | --- | --- | --- | --- | --- | --- |
| 1 | 31-35 | 24.3 | 196 | Posterior | 266 | 2730 | 10.8 |
| 2 | 16-20 | 29 | 169 | Anterior | 280 | 3289 | 19.7 |
| 3 | 26-30 | 36.9 | 176 | Anterior | 292 | 2900 | 1.3* |
| 4 | 27-30 | 37.9 | 172 | Posterior | 265 | 2900 | 17 |
| 5 | 36-40 | 28.3 | 195 | Posterior | 270 | 3420 | 59.4 |
| 6 | 21-25 | 25 | 169 | Anterior | 282 | 3674 | 41 |
| 7 | 36-40 | 18.1 | 178 | Posterior | 273 | 3682 | 81 |
| 8 | 26-30 | 29.7 | 174 | Posterior | 236 | 2660 | 98 |
| 9 | 26-30 | 26.8 | 170 | Anterior | 275 | 2500 | 1.4* |
| 10 | 31-35 | 17.4 | 176 | Posterior | 286 | 2950 | 12 |
| 11 | 26-30 | 21.3 | 106 | Posterior | 287 | 3106 | 15 |
| 12 | 31-35 | 21.5 | 194 | Anterior | 272 | 2830 | 7 |

Subjects were scanned using a 1.5T Philips Achieva MRI scanner (Philips Medical Systems Best, NL) whilst supine with a left lateral tilt, to reduce aortocaval compression by the gravid uterus. A cardiac-receiver coil was placed on the abdomen, covering the entire uterus. Non-rebreathing masks (Intersurgical, Wokingham, UK) delivered medical air or 100% oxygen at a flow rate of 15 L/min to the subjects and respiratory triggering was used to minimise motion from maternal breathing.

T_2_-weighted structural scans were used to determine the position of the placenta and plan the OE-MRI slices. Two inversion recovery OE-MRI protocols were performed: a 3D turbo-spin echo with half Fourier acquisition (3D-HASTE) for full placental coverage and a 2D turbo-spin echo (2D-TSE), as used in previous studies(2,3). The single 2D slice positioned perpendicular to the placenta at the level of the cord insertion. The orientation of the 3D-HASTE matched that of the 2D-TSE. For both OE-MRI protocols a T_1_ map was calculated using a set of inversion recovery images, followed by a dynamic series of T_1_-weighted images during which the gas supply to the subject is switched from medical air (21%) to 100% oxygen in order to determine ΔR_1_.

### 3D OE-MRI protocol

The majority of the data were acquired as follows: T_1_ mapping data for the 3D-HASTE acquisitions consisted of 5 separate acquisitions with inversion times (TI): 50, 300, 1100, 2000 ms and no inversion pulse (≈TR of 8 s, dependant on triggering), TE of 9 ms, 450 × 450 mm^2^ FOV, matrix size 176 × 176, voxel size 2.56 × 2.56 mm^2^, slice thickness 10 mm and 48 slices. Respiratory-triggered T_1_-weighted images were acquired dynamically at 40 time points with a temporal resolution of ∼8 s (triggered off of every other breath), a TI of 1400 ms, with matching TE, FOV, matrix size, voxel size, slice thickness and number of slices. On the 10 dynamic, medical air was switched to 100% oxygen.

On completion of the 3D dynamic acquisition, the supply was switched back to medical air and a 2-minute interval was observed to return the oxygen levels to baseline.

#### Updated protocol

In 2 data sets (subjects 11 and 12) the first TI, matrix size and voxel size were changed to match the 2D OE-MRI protocol below (60 ms, 128 × 128, 3.52 × 3.52 mm^2^).

### 2D OE-MRI protocol

T_1_ maps for the 2D-TSE acquisitions consisted of 5 inversions times (TI): 60, 300, 1100, 2000 ms and no TI. TR = TI + 8000 ms, TE of 5.4 ms, 450 × 450 mm^2^ FOV, matrix size 128 × 128, voxel size 3.52 × 3.52 mm^2^ and slice thickness 10 mm. Respiratory-triggered T_1_-weighted images were acquired dynamically at 40 time points with a temporal resolution of ∼9.4 s and a TI of 1400 ms. On the 10^th^ dynamic, medical air was switched to 100% oxygen.

### Analysis

Data were analysed using in-house Python code. An ROI was defined on the TI = 300 ms image, which incorporated the entire placenta as observed on each slice. Care was taken on the fetal side of the placenta and at each end to exclude areas likely to be affected by motion. A T_1_ map was created by fitting a 3-parameter inversion recovery model to the data on a voxel-wise basis. The baseline/on air T_1_ (T_1b_) was used to calculate the change in R_1_ (ΔR_1_) for each dynamic in the T_1_-weighted series, as described elsewhere(1). The single slice from the 2D OE-MRI (ss-2D) was compared with a single matched slice from the 3D volume (ss-3D) and the entire volume (vol-3D). The slice was matched by using the 3D slice with the closest slice location, according to the DICOM header and checked visually.

Voxel-wise ΔR_1_ was calculated by taking the mean of the last 20 dynamics during the oxygen plateau and plotted as maps for both 2D and 3D data. Median and inter-quartile range (IQR) across the ROI of ΔR_1_ and T_1b_ were calculated for each subject, to limit the influence of outliers in the voxel-wise maps. The correlation was calculated for median and IQR ΔR_1_ and T_1b_ against gestational age at the scan, as in (1–3). The significance of the correlation was measured by using the Pearson correlation coefficient and presented as r and P values.

Bland–Altman plots were produced to quantitatively assess the agreement between the two methods, with 95% confidence intervals (limits of agreement) calculated as the mean difference ± (1.96 × standard deviation of the difference). The ROIs for subjects identified as outliers on these plots were inspected visually on the dynamic data.

Example videos of the 3D ΔR_1_ maps can be found in the online supplementary material. Two women who did not meet the outcome criteria for assignment to normal group (IBR <5^th^ percentile) were removed from the gestational correlation analysis. All 12 subjects remained in all other comparisons.

## Results

In Figure 1 the subject-average ΔR_1_ is plotted against dynamic number for the ss-2D data and the vol-3D data. For both, the signal increases from baseline after the oxygen concentration is increased (10^th^ dynamic) and plateaus by the 20^th^ dynamic image. The ss-2D and vol-3D curves plateaus around the same ΔR_1_ with a similar standard deviation on each dynamic ΔR_1_.

**Figure 1.**
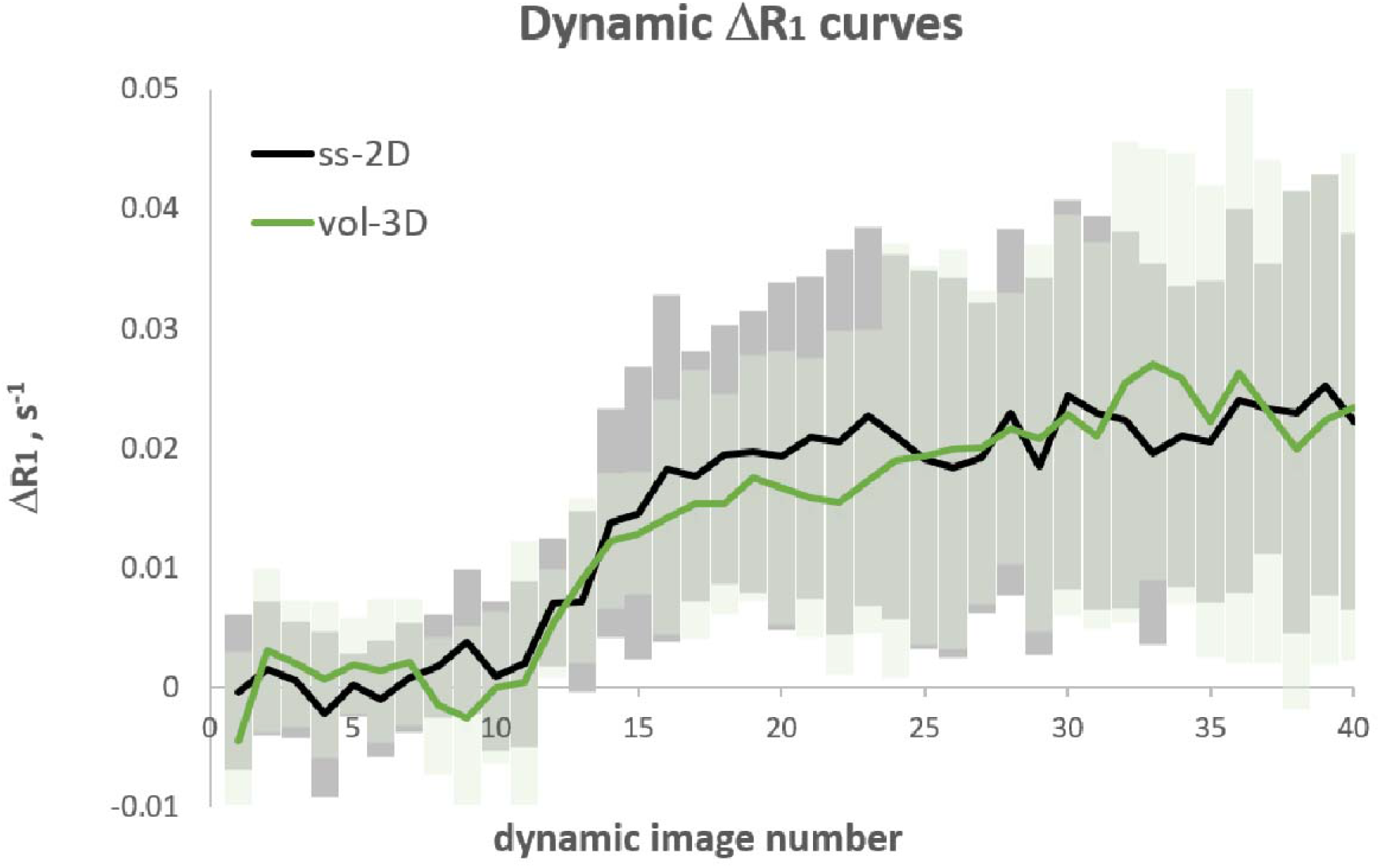
Mean (line) and standard deviation of the dynamic ΔR_1_ curves against dynamic image number for ss-2D (grey) compared with vol-3D (green).

Table 2 shows the group average median ΔR_1_ for ss-2D (0.020 s^-1^), ss-3D (0.023 s^-1^) and vol-3D (0.022 s^-1^) and that ss-3D (p = 0.58) and vol-3D (p = 0.70) do not differ significantly from ss-2D when compared using a two-tailed paired t-test. However, median T_1b_ for ss-2D was higher (1603 ms) than T_1b_ for ss-3D (1540 ms, p=0.07) and significantly higher than vol-3D (1515 ms, p = 0.02), when compared using a two-tailed paired t-test. Correlation of median ΔR_1_ with gestation age at scan for the normal group (N = 10) revealed no trend with gestational age for ss-2D (p = 0.99), ss-3D (p = 0.14) and vol-3D (p = 0.24).

**Table 2.** Group average median and IQR ΔR_1_ and T_1b_ values (12 subjects) and group average median and IQR ΔR_1_ and T_1b_ values against gestational age at scan (10 subjects) for ss-2D, ss-3D and vol-3D.

|  | ss-2D | ss-3D | vol-3D |
| --- | --- | --- | --- |
| **median $\Delta R_1$, s<sup>-1</sup>** | 0.020<br>(0.004-0.038) | 0.023<br>(-0.006-0.047) | 0.022<br>(-0.003-0.048) |
| **IQR $\Delta R_1$, s<sup>-1</sup>** | 0.024<br>(0.012-0.048) | 0.025<br>(0.015-0.045) | 0.027<br>(0.015-0.038) |
| **median $\Delta R_1$ – gestation** | r 0.00<br>p 0.99 | r -0.25<br>p 0.14 | r -0.17<br>p 0.24 |
| **IQR $\Delta R_1$ – gestation** | r -0.18<br>p 0.23 | **r -0.45**<br>**p 0.03** | **r -0.64**<br>**p 0.005** |
| **median T<sub>1b</sub>, ms** | 1603<br>(1305-1679) | 1540<br>(1418-1693) | 1515<br>(1374-1723) |
| **IQR T<sub>1b</sub>, ms** | 177<br>(86-275) | 196<br>(122-338) | 217<br>(152-340) |
| **median T<sub>1b</sub> – gestation** | r 0.22<br>p 0.17 | r 0.11<br>p 0.77 | r 0.00<br>p 0.99 |
| **IQR T<sub>1b</sub> – gestation** | r 0.19<br>p 0.20 | **r -0.39**<br>**p 0.05** | **r -0.45**<br>**p 0.03** |

Table 2 also shows the inter-quartile range (IQR) of ΔR_1_, a measure of heterogeneity. The ss-2D values are not significantly different from ss-3D (p = 0.94) or vol-3D (p = 0.36) when compared using a two-tailed paired t-test. Correlation of IQR ΔR_1_ with gestation age at scan revealed lower values at longer gestation for the 3D data, ss-3D (p = 0.03) and vol-3D (p = 0.005).

A Bland–Altman plot shown in Figure 2 was used to investigate the agreement between ΔR_1_ for ss-2D with ss-3D and vol-3D for the individual subjects. The plots show no outliers for ss-2D and ss-3D or ss-2D and vol-3D. The 2D - 3D difference in ΔR_1_ is a smaller magnitude than the mean ΔR_1_. The average difference appears to decrease slightly as ΔR_1_ increases. Figure 3 show the Bland–Altman plots comparing the T_1b_ values acquired from the 2D and 3D scans. The plots show a single outlier for ss-2D and ss-3D and no outliers for ss-2D and vol-3D. The mean difference shows the trend towards lower T_1b_ values for ss-3D and vol-3D, compared with ss-2D.

**Figure 2.**
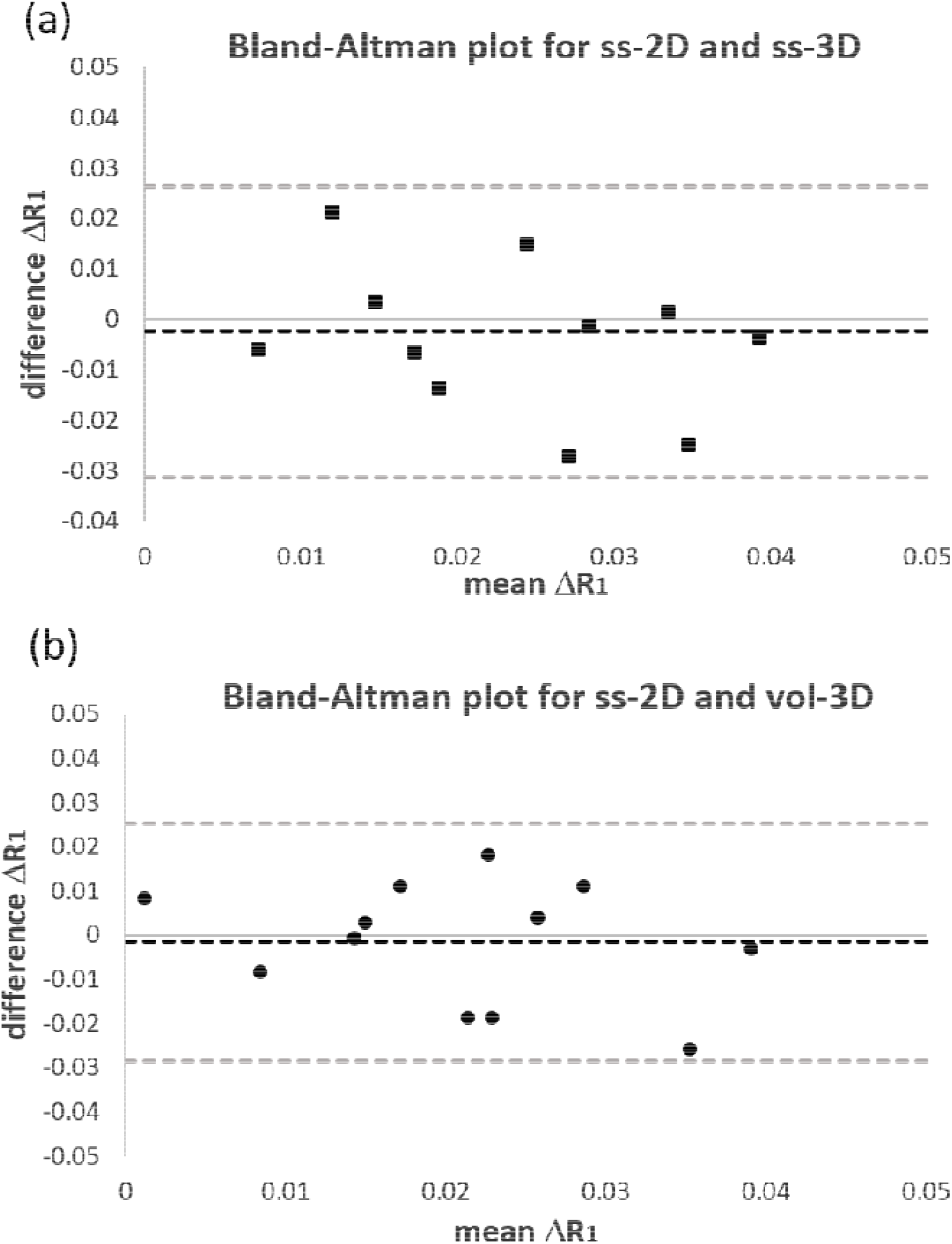
Bland–Altman plots of mean ΔR_1_ against the difference between 2D and 3D ΔR_1_ for (a) ss-2D and ss-3D and (b) ss-2D and vol-3D. The black dotted line represents the mean difference and the grey dotted lines the limits of agreement (95% confidence interval).

**Figure 3.**
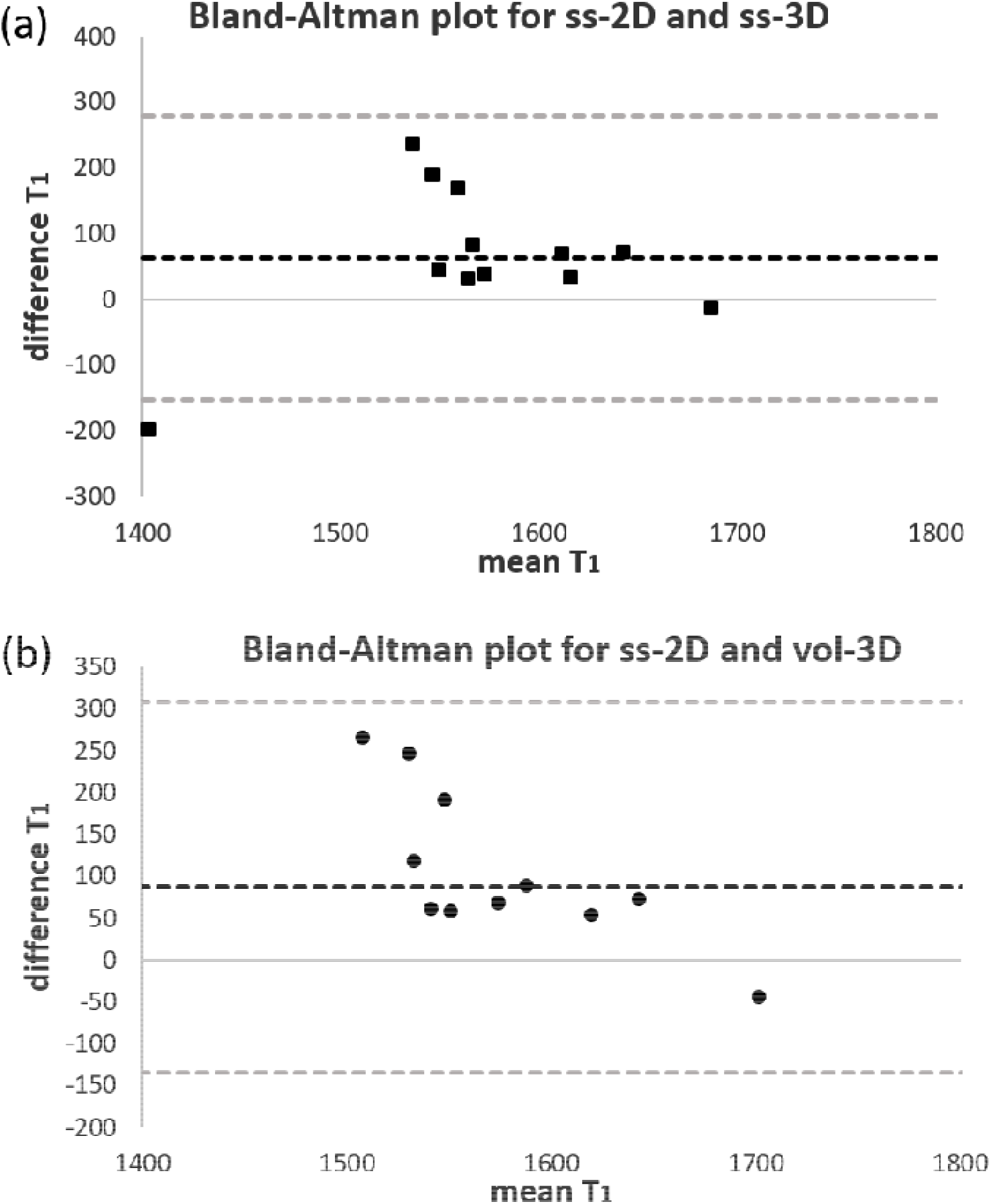
Bland–Altman plots of mean T_1b_ against the difference between 2D and 3D T_1b_ for (a) ss-2D and ss-3D and (b) ss-2D and vol-3D. The black dotted line represents the mean difference and the grey dotted lines the limits of agreement (95% confidence interval).

Figure 4 shows maps comparing the voxel-wise ΔR_1_ for ss-2D and a matching slice from the 3D volume for an example anterior (Figures 4a and 4b) and posterior (Figures 4c and 4d) placenta. The 3D images exhibit similar features to the 2D maps for both of the placenta, however there are amplitude differences for the posterior placenta. Qualitatively, the ss-3D and vol-3D maps are less noisy and more blurred.

**Figure 4.**
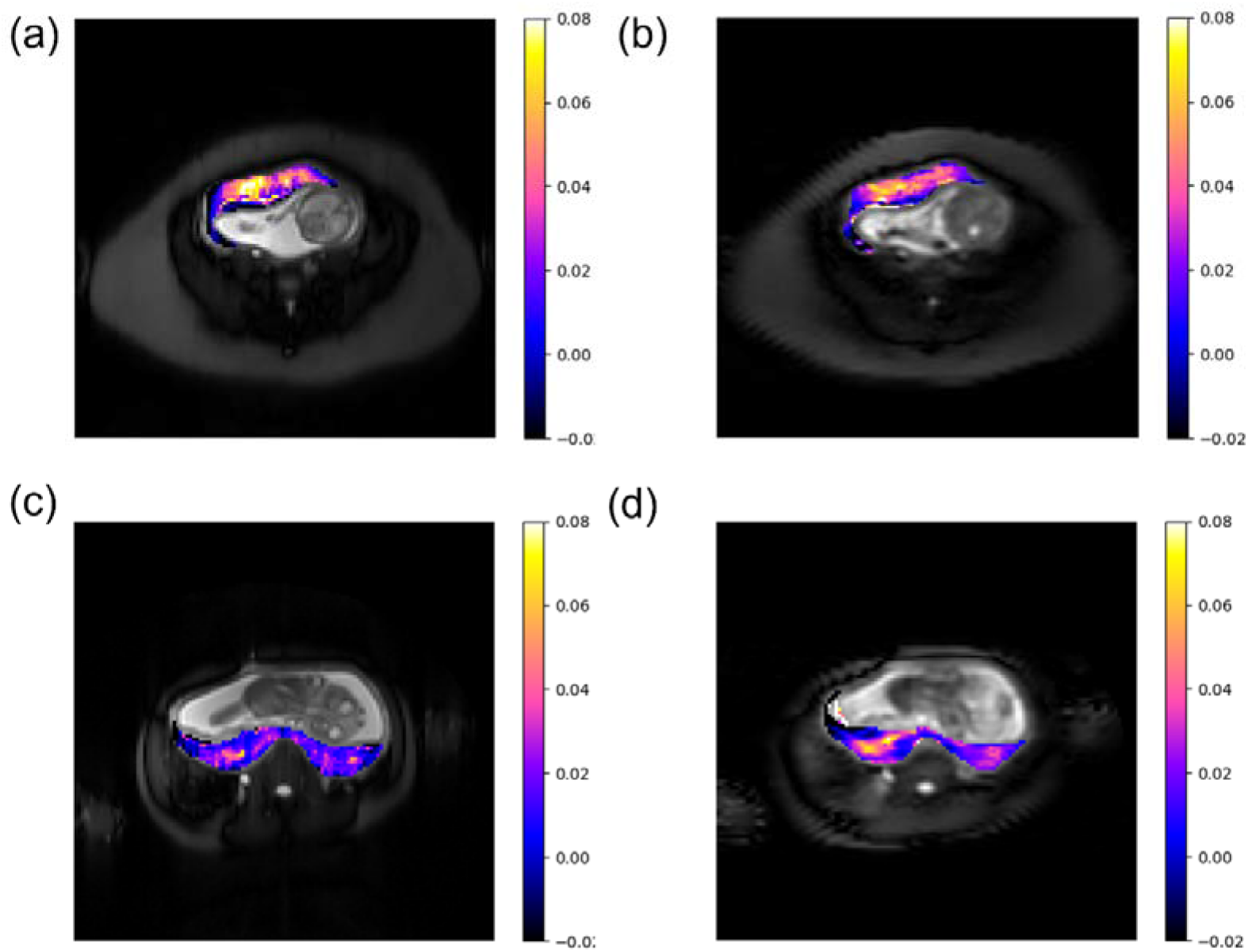
*Top*: ΔR_1_ (s^-1^) maps of an example anterior placenta for (a) ss-2D and a matching (b) ss-3D; *Bottom*: ΔR_1_ (s^-1^) maps of an example posterior placenta for (a) ss-2D and a matching (b) ss-3D.

## Discussion

This study shows that it is feasible to acquire whole placental volume OE-MRI data on a Philips 1.5 T MRI scanner. All scans were well tolerated by the pregnant subjects, even though increasing the coverage to 3D required the use of a cardiac-receiver coil in addition to the non-rebreathing mask.

There were no significant differences between median ΔR_1_ for the original single slice 2D protocol (ss-2D) compared with a matched single slice from the 3D data (ss-3D) or the full 3D volume (vol-3D), the individual difference between the 2D and 3D data was on the same order of magnitude as the group average and showed some scatter. All subjects fall within the limit of agreement (95% CI) on the ΔR_1_ Bland–Altman plots in Figure 2. Only subject 1 falls outside the limit for the ss-2D v ss-3D comparison of T_1b_ (Figure 3). This subject has T_1b_ in the normal range for ss-3D and vol-3D, but the 2D data shows a low median T_1b_ (ss-2D 1305 ms compared to an average of 1603 ms).

Previous studies have shown a negative correlation of ΔR_1_ (1–3) and T_1b_ (11,12) with gestational age at scan. Here no correlation was significant in this study. However, this study included a relatively small group of subjects (10 compared with between 9(2) and 41(12)) and much smaller gestational window (169-196 days compared with up to 145-2671 days(2)). The study cohort was primarily chosen to allow the quality of the data acquired using the 3D protocol to be compared with that of the original 2D protocol, not to test for correlation with gestation. However, a negative trend in IQR ΔR_1_ and IQR T_1b_ was apparent for 3D data (Table 2) corresponding to a reduction in heterogeneity with gestation. The two subjects that were removed from correlation of median R_1_ with gestational age at scan, due to an IBR < 5%, had relatively low or abnormal ΔR_1_. This is in qualitative agreement with the results of (3) (see maps in supplementary material – subjects 5 and 9), however the number of subjects is too small to perform a quantitative comparison.

Figure 4 shows that the underlying images are of poorer quality for the 3D data (b and d) than for the 2D data (a and c), with more artefact and significant T_2_ blurring due to the long echo train and/or undersampling. However, similar features are observed for both the 2D and 3D ΔR_1_ maps. ΔR_1_ appears to be greater in the intermediate section of the placenta, with a decreasing gradient towards both the fetal and maternal sides. The 3D ΔR_1_ video maps in supplemental material show that the gradient is observable throughout the extent of the placenta. The higher values of ΔR_1_ appear in the intermediate region where the villous tree exchanges oxygen between the maternal and fetal blood(13). This gradient may be visible as a result of the higher signal-to-noise ratio of the 3D volume acquisition compared with the 2D, but it may be artificially enhanced by partial volume effects at the boundary between the placenta and the surrounding tissue due to image blurring. No statistically significant difference in the heterogeneity measurement IQR between the 2D and the 3D ΔR_1_ data was observed, however the correlation of IQR with gestational age shows a statistically significant decrease for both the ss-3D (p = 0.03) and vol-3D (p = 0.005), but not for ss-2D. This may be a result of an overall lower ΔR_1_ at later gestation leading to a smaller range or it may reflect a real increase in the homogeneity of oxygenation with increasing gestation to which the 3D data is sensitive.

There are number of limitations of the study. Firstly, the ROI was drawn on the TI = 300ms and applied to the dynamic series of data. The use of the 300ms data for region definition allowed the placenta to be differentiated more clearly, however any gross motion between this scan and the dynamic acquisition used to calculate ΔR_1_ could have a detrimental effect on the resulting maps and statistics. Secondly, unlike in previous OE-MRI studies (2,3) where the ROI has been trimmed to only include voxels where placental tissue was present throughout the whole dynamic, the placental volume for each slice was included without trimming. Care was taken on the fetal side of the placenta to not include approximately one voxel around the amniotic sac to reduce the effects of fetal motion. Any significant motion was accepted as noise on the ΔR_1_ curve, with the intention of not unnecessarily removing placental tissue and biasing the analysis by drawing small ROIs. Thirdly, the 3D protocol was updated during the study to match the dimensions of the 2D protocol. This does not appear to have any significant effect on the quality of the data or the resulting maps, but the number of scans with the updated protocol is too small to test for any bias. Finally, the slice matching to obtain the ss-3D data was performed by using the slice location from the DICOM header. This does not take into account gross motion between scans and is only an approximate due to even number of slices in 3D.

The aetiology of FGR is variable with specific phenotypes due to chromosomal, infectious, vascular disease and inflammatory-related causes. It is hoped that whole volume OE-MRI acquisition may allow for in utero placental phenotyping in FGR, thereby improving disease stratification, therapeutic intervention and outcome management. FGR placentas are known to have poor peripheral vascularisation and visible hypo-vascular regions(6) Other gross lesions evident in maternal vascular malperfusion include placental hypoplasia, infarction and altered villous development(14). These abnormalities are likely to lead to heterogeneity in the oxygenation across the placental volume that will be more readily observable when imaging the whole placental volume as opposed to a single central slice. A reliable measure of placental oxygenation across the entire placenta is a welcome addition to the MRI toolbox and could be utilised with other clinical markers and imaging to aid identification of the placental phenotype of FGR and improve fetal outcomes. Here we show that it is possible to obtain a measure of oxygenation via ΔR_1_ across the entire placenta. The 3D OE-MRI measurement is sensitive to changes in placental oxygenation and allow mapping of the whole placental volume.

## Data Availability

All imaging data produced in the present study are available upon reasonable request to the authors. All code and summary data are availaible in the supplementary material.

## Acknowledgments

This project was funded by Tommy’s the baby charity. MRI scans were undertaken within Wolfson Molecular Imaging Centre.

## Notes

### Competing Interest Statement

The authors have declared no competing interest.

### Funding Statement

This study is funded by Tommy's.

### Author Declarations

North West Greater Manchester East/West Research Ethics committee gave ethical approval for this work. REC:09/H1013/77 and 14/NW/0195.

