## supplementary material for "Comparison of 2D and 3D oxygen-enhanced MRI of the placenta"

### Slide 1
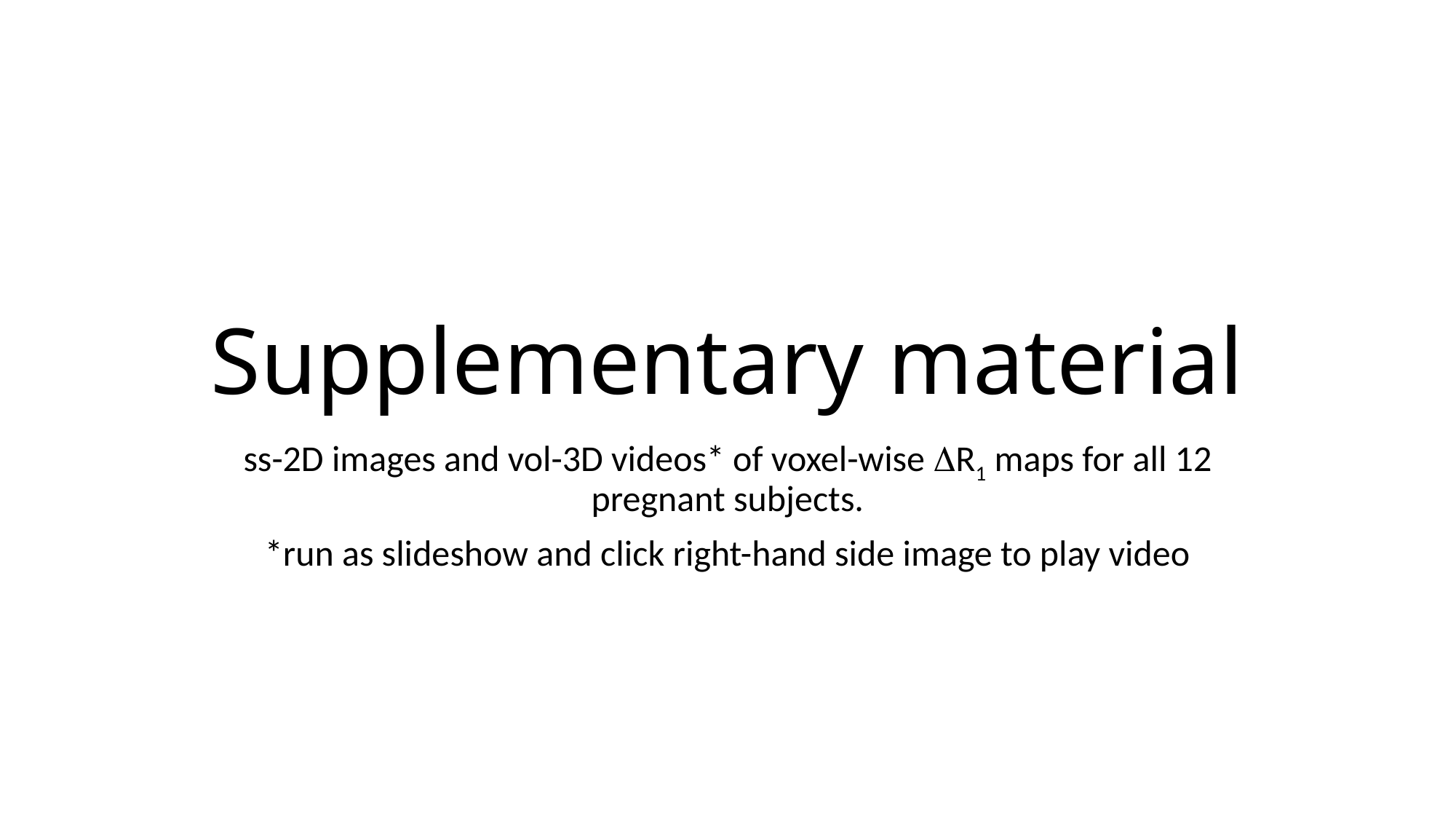

#### Supplementary material
ss-2D images and vol-3D videos* of voxel-wise ΔR_1_ maps for all 12 pregnant subjects.
*run as slideshow and click right-hand side image to play video

### Slide 2
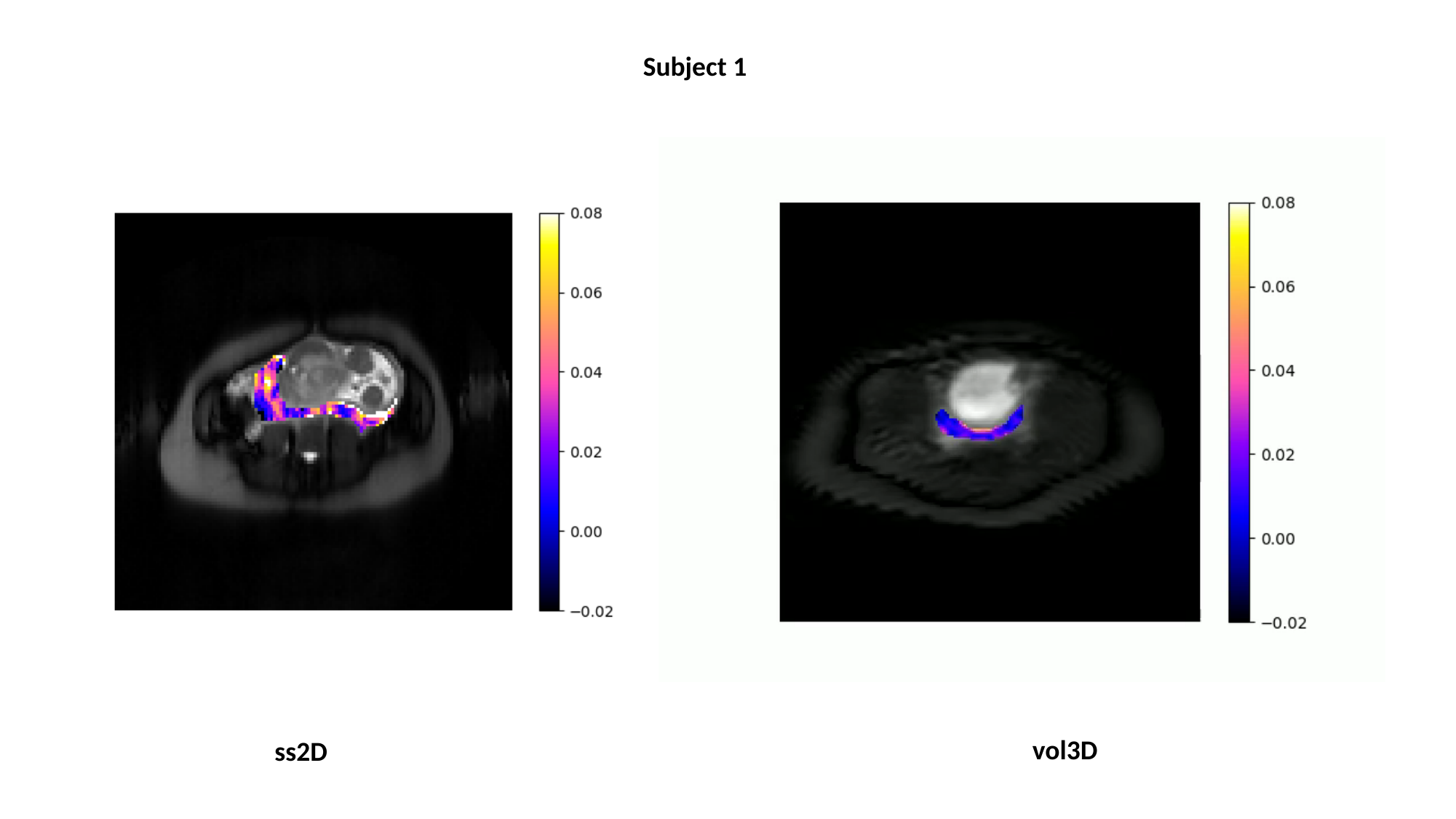

Subject 1
vol3D
ss2D

### Slide 3
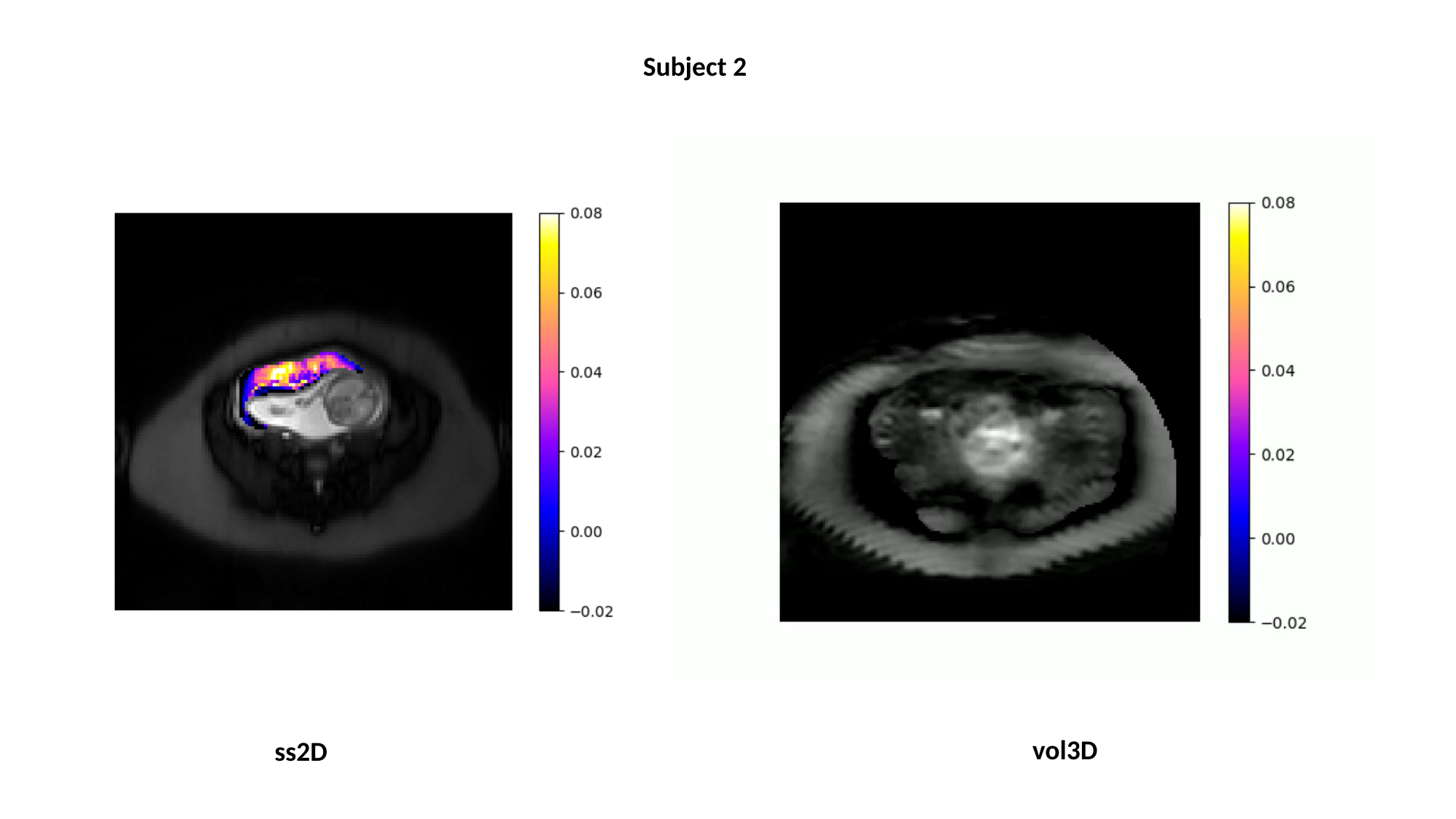

Subject 2
vol3D
ss2D

### Slide 4
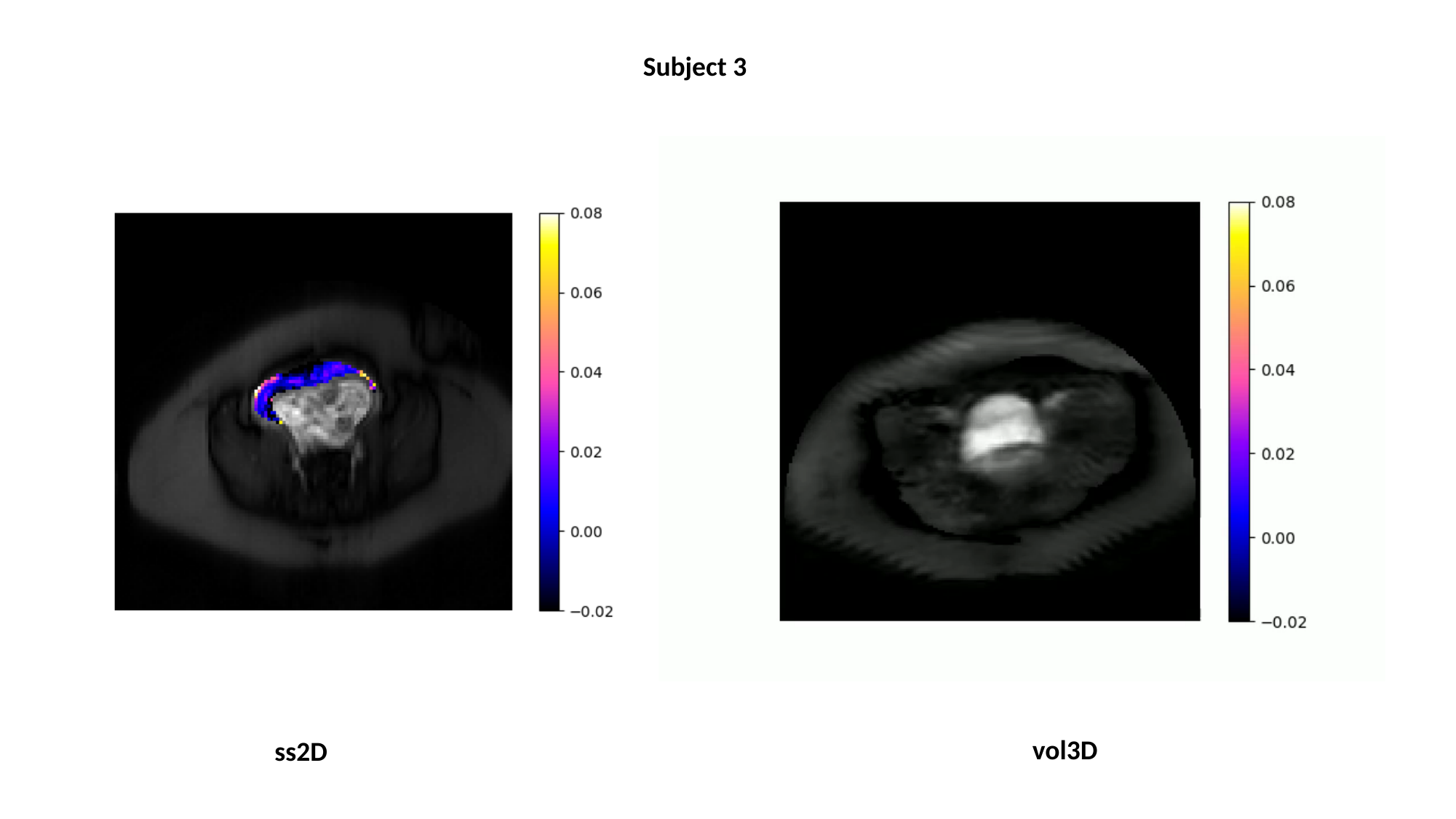

Subject 3
vol3D
ss2D

### Slide 5
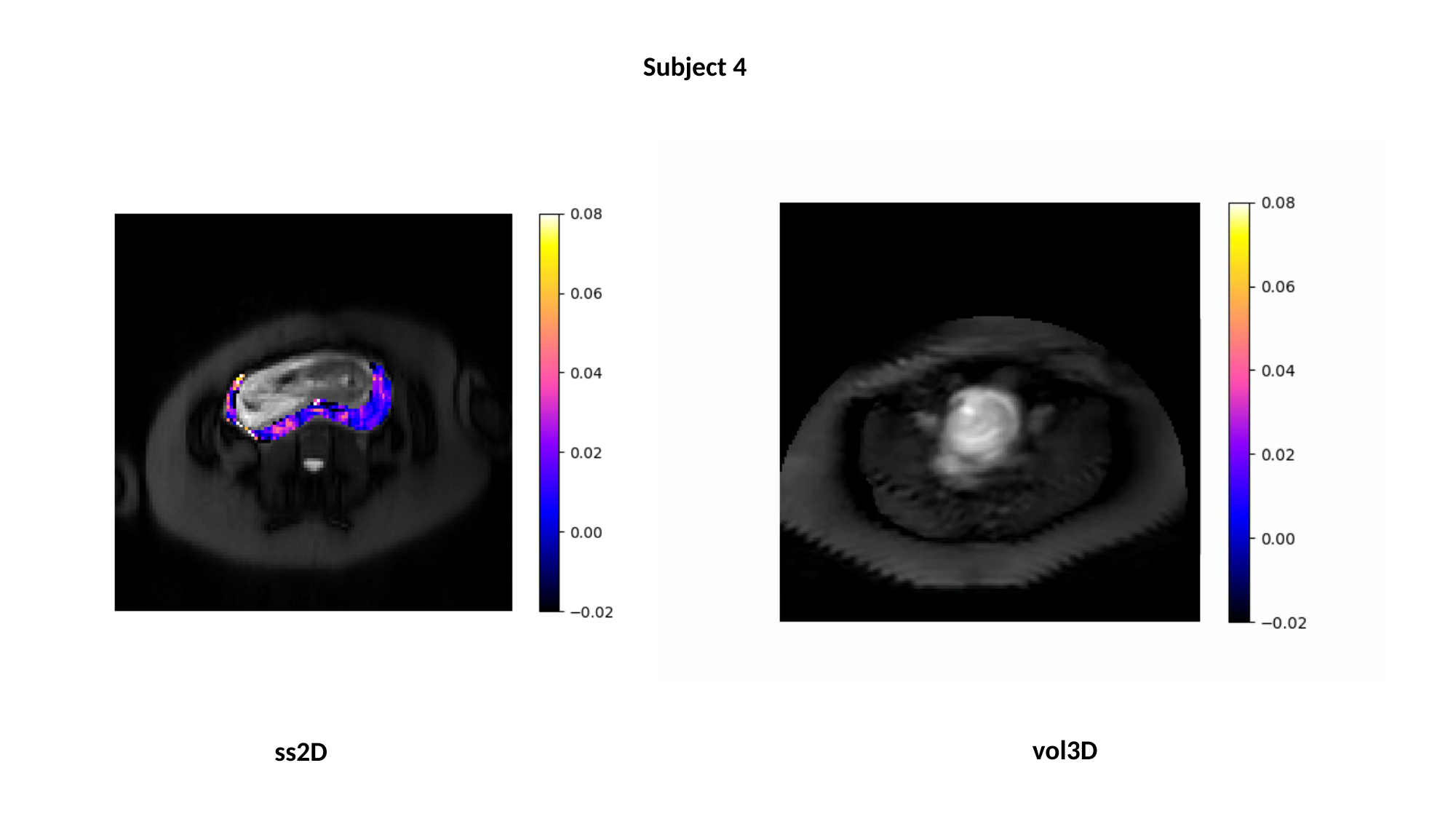

Subject 4
vol3D
ss2D

### Slide 6
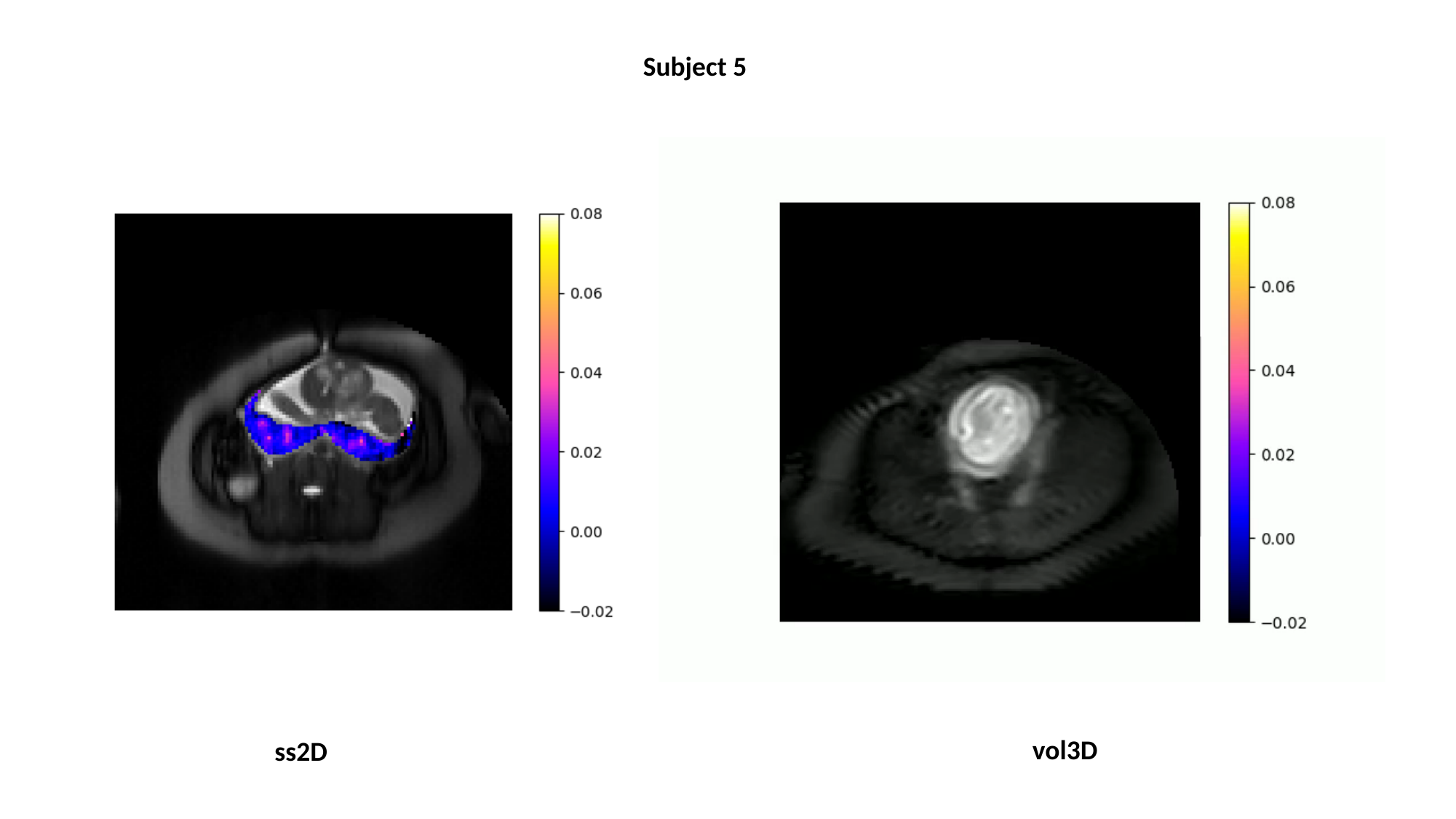

Subject 5
vol3D
ss2D

### Slide 7
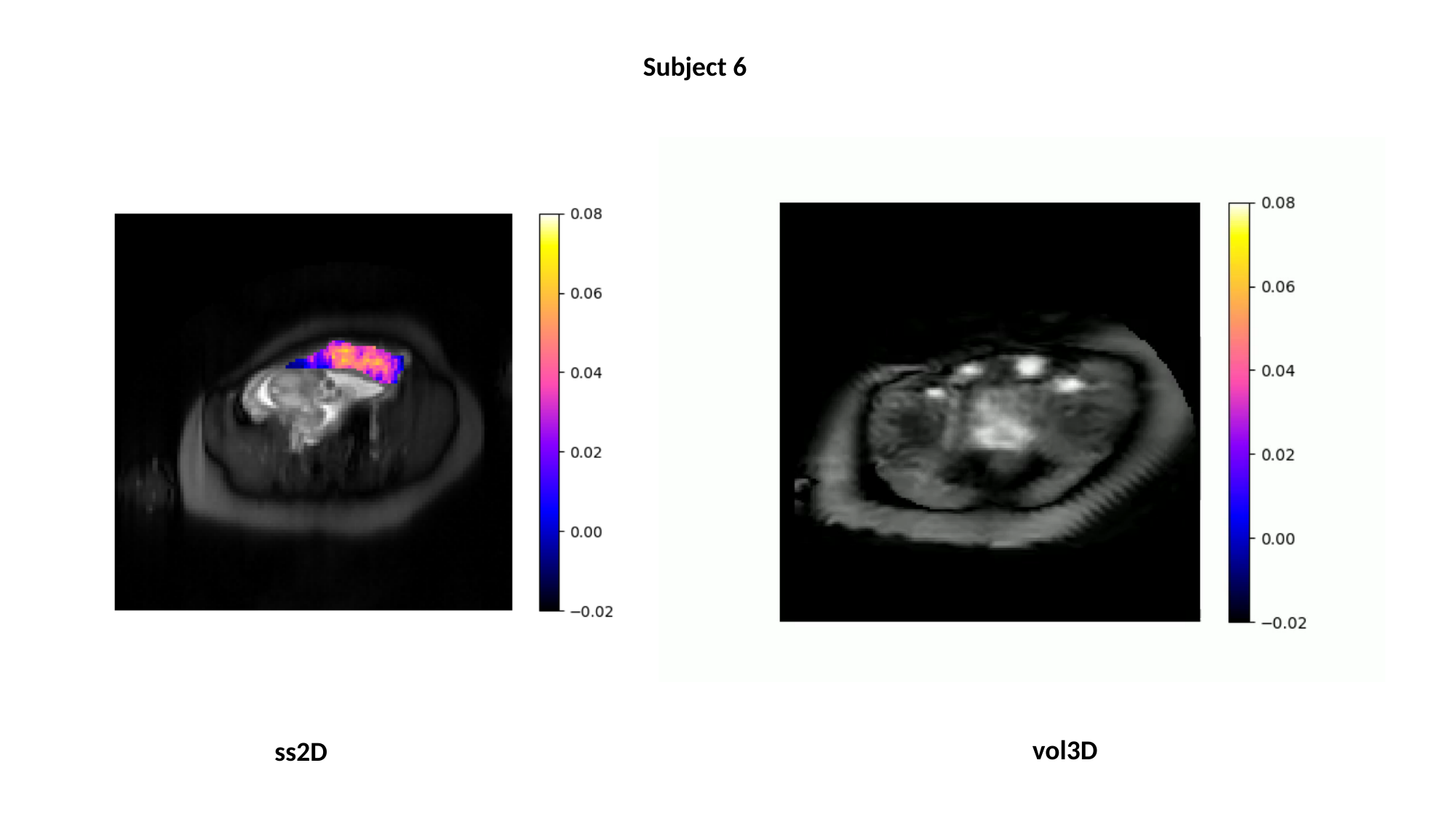

Subject 6
vol3D
ss2D

### Slide 8
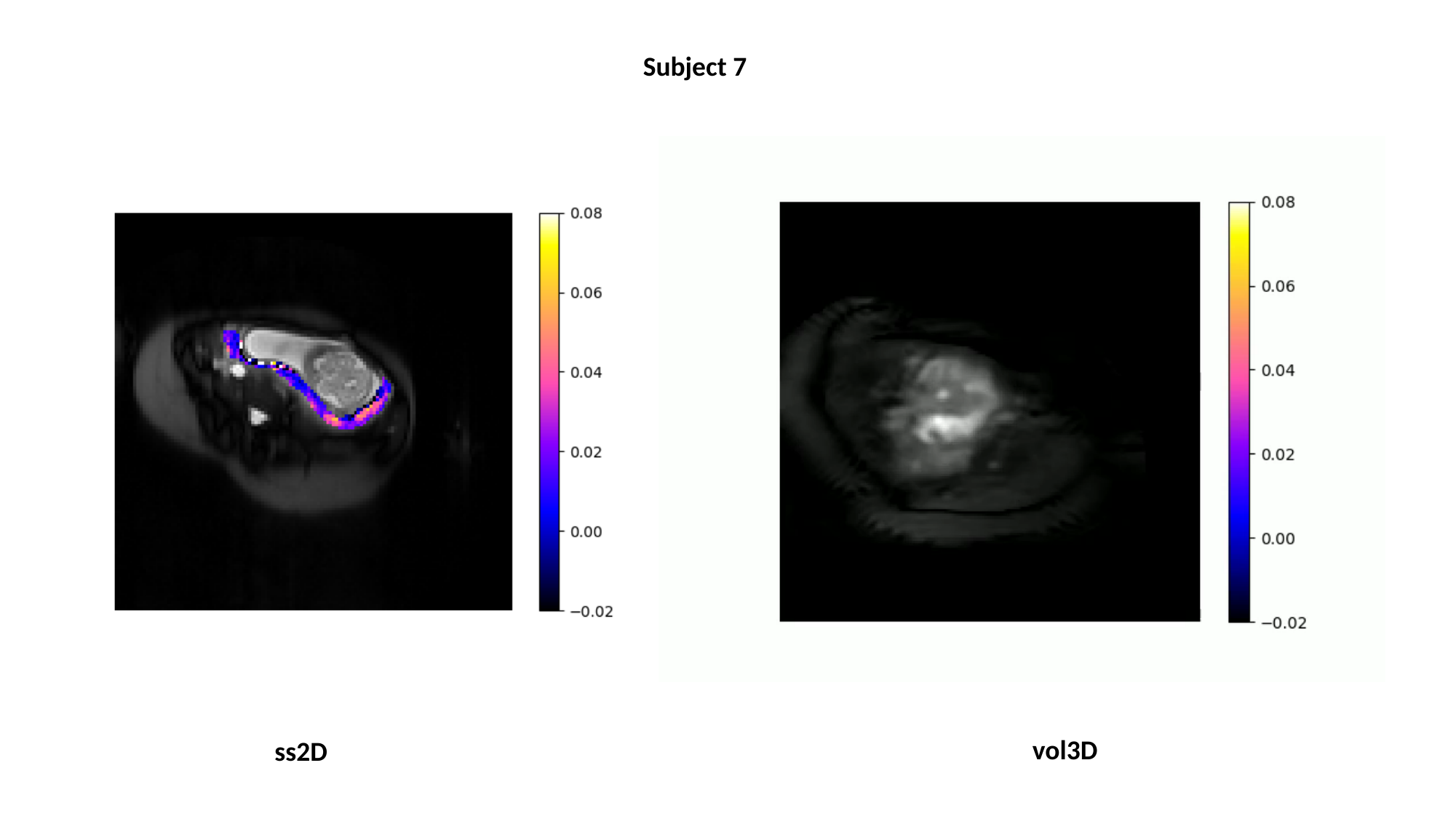

Subject 7
vol3D
ss2D

### Slide 9
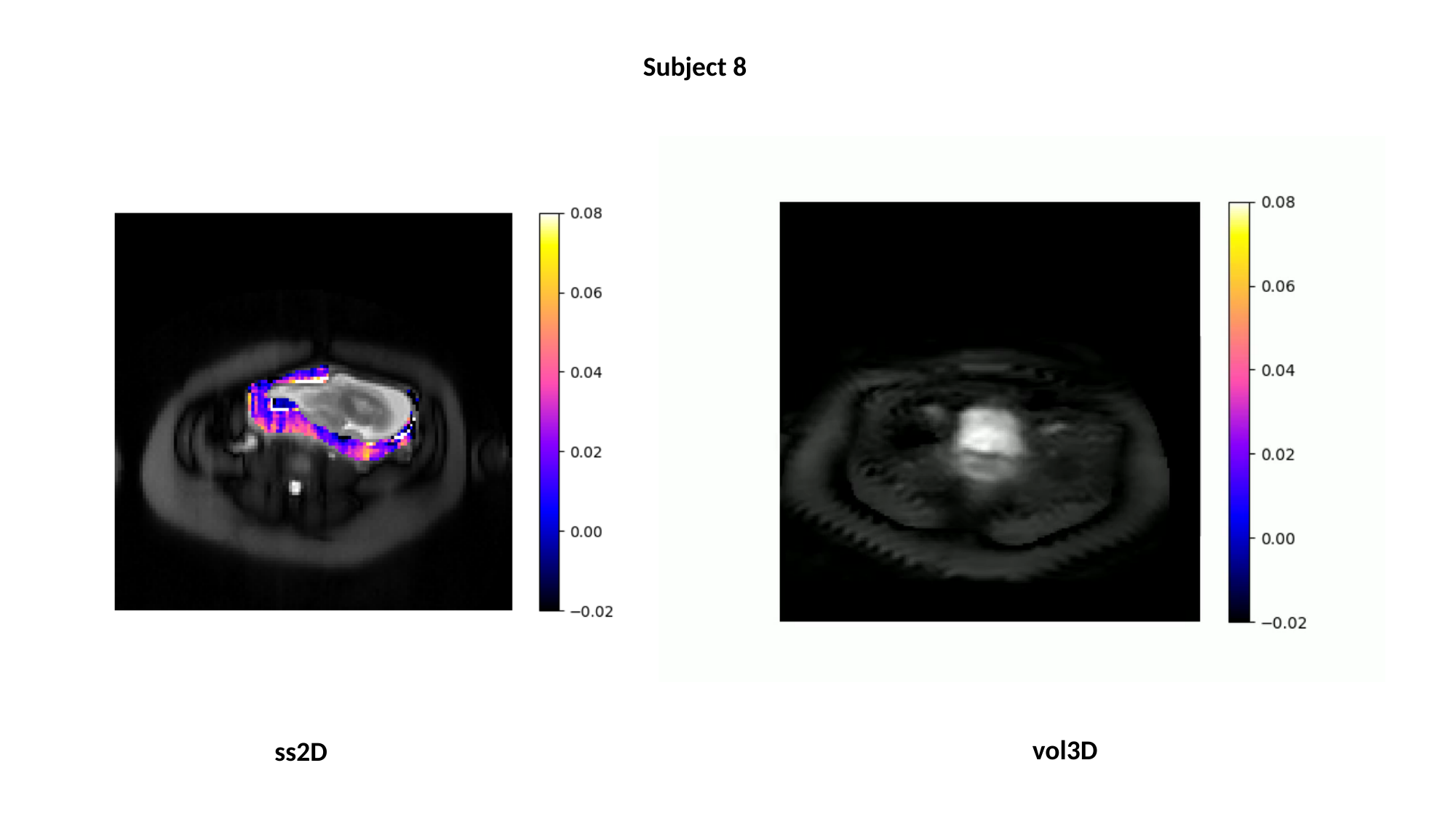

Subject 8
vol3D
ss2D

### Slide 10
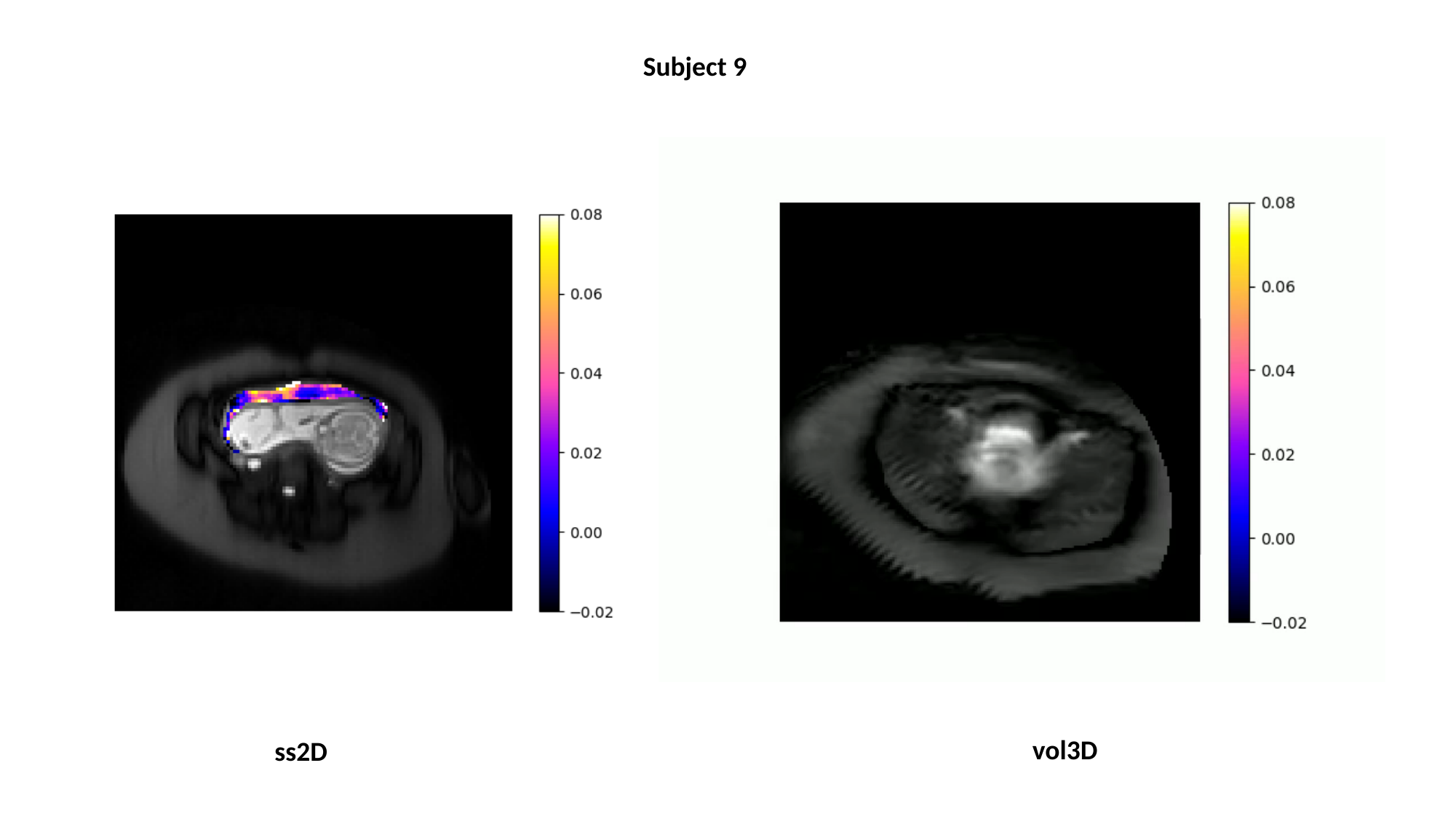

Subject 9
vol3D
ss2D

### Slide 11
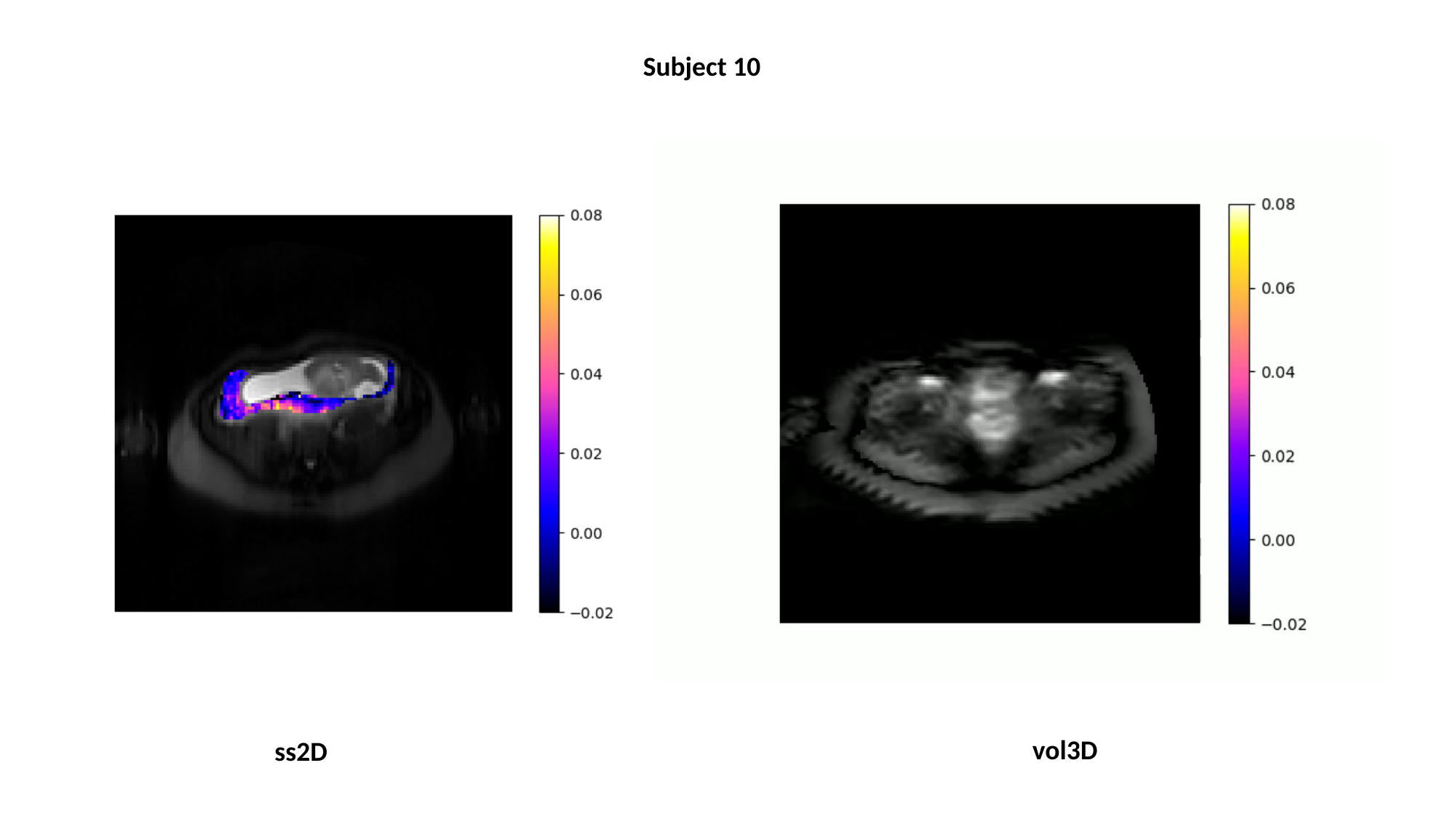

Subject 10
vol3D
ss2D

### Slide 12
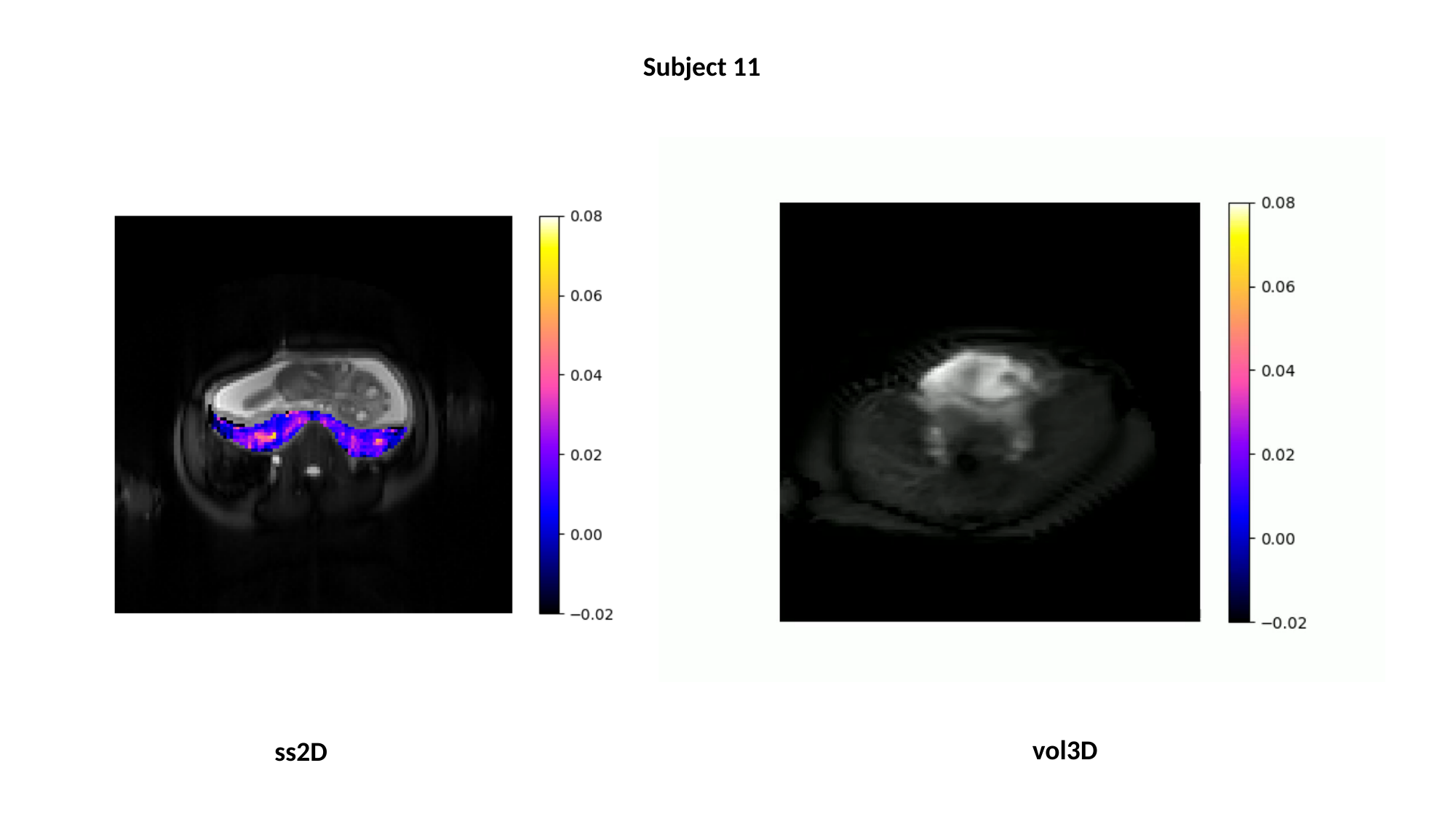

Subject 11
vol3D
ss2D

### Slide 13
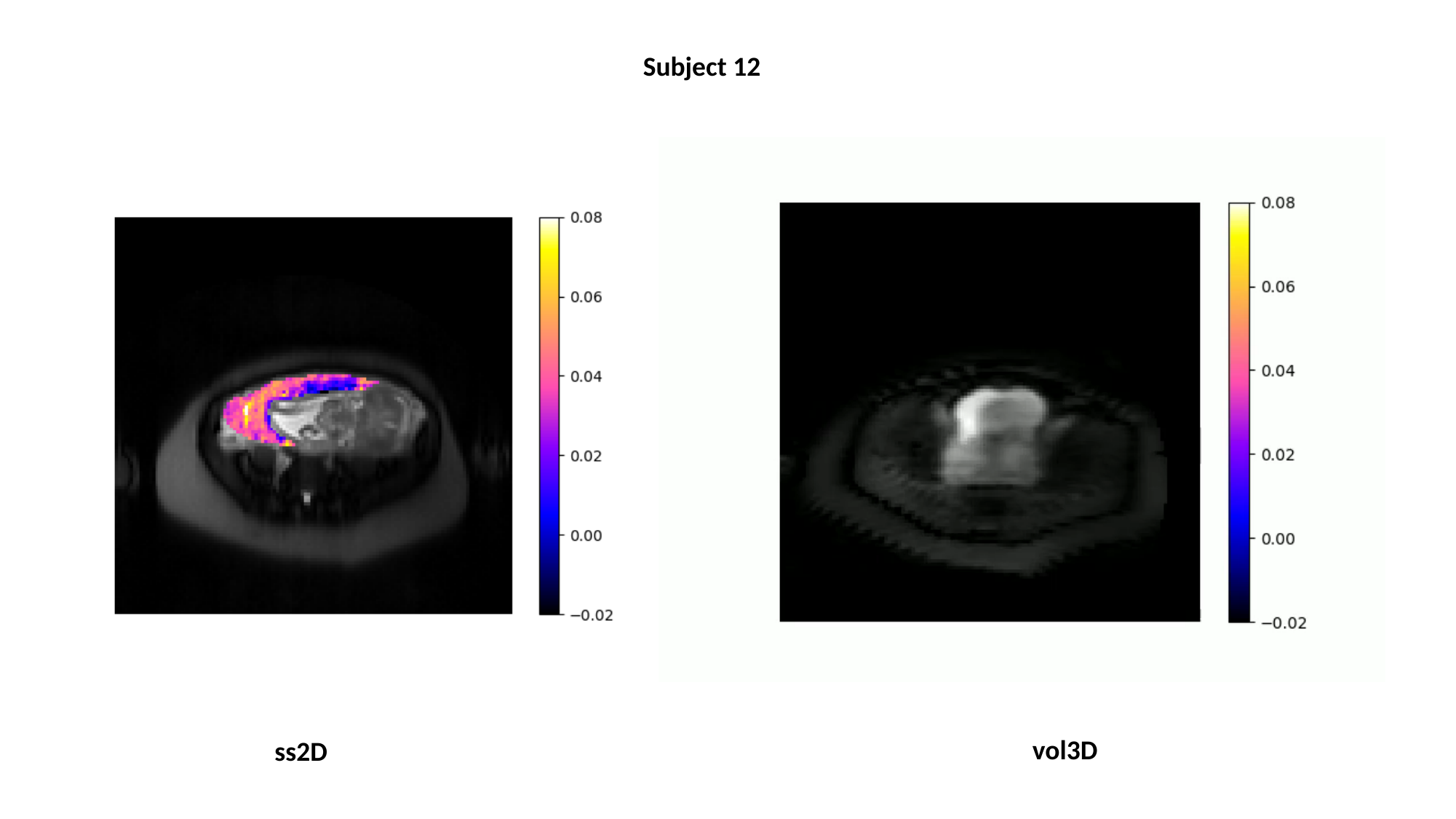

Subject 12
vol3D
ss2D
